# Self-reported work activities, mucus membrane symptoms, and respiratory health outcomes among an industrial hog operation worker cohort, North Carolina, USA

**DOI:** 10.1101/2020.09.29.20203893

**Authors:** Vanessa R. Coffman, Devon J. Hall, Nora Pisanic, David C. Love, Maya Nadimpalli, Meredith McCormack, Marie Diener-West, Meghan F. Davis, Christopher D. Heaney

## Abstract

**Background:** Respiratory disease among industrial hog operation (IHO) workers is well documented; however, it remains unclear whether specific work activities are more harmful and if personal protective equipment (PPE), as used by workers, can reduce adverse health outcomes.

**Objectives:** To assess the relationship between self-reported IHO work activities and PPE use with mucus membrane and respiratory health symptoms in an occupational cohort.

**Methods:** IHO workers (*n*=103) completed baseline and up to eight bi-weekly (*i.e*., every two weeks) study visits. Workers reported typical (baseline) and transient (bi-weekly) work activities, PPE use, and physical health symptoms. Baseline and longitudinal associations between work activities and health outcomes were assessed using generalized logistic and fixed-effects logistic regression models, respectively.

**Results:** At baseline, reports of ever versus never drawing pig blood, applying pesticides, and increasing years worked at any IHO were positively associated with reports of eye, nose, and/or throat irritation. Over time, transient exposures, including those associated with dustiness in barns, cleaning of barns, and pig contact were associated with increased odds of symptoms including sneezing, headache, and eye or nose irritation, particularly in the highest categories of exposure. When PPE was used, workers had decreased odds of symptoms interfering with sleep (odds ratio (OR): 0.1; 95% confidence interval (CI): 0.01, 0.8), sneezing (OR: 0.1; 95% CI: 0.01, 1.0), and eye or nose irritation (OR: 0.1; 95% CI: 0.02, 0.9). Similarly, when they washed their hands ≥8 times per shift (the median) versus less frequently, workers had decreased odds of any respiratory symptom (OR: 0.3; 95% CI: 0.1, 0.8).

**Discussion:** In this healthy volunteer IHO worker population, increasingly unfavorable work activities were associated with self-reported mucus membrane and respiratory health outcomes. Strong protective associations were seen between PPE use and handwashing and the odds of symptoms, warranting further investigation in intervention studies.

## Introduction

Industrial hog operations (IHOs) pose a respiratory health risk to workers (Casey et al. 2015; Cole et al. 2000; Heederik et al. 2007; Kirkhorn et al. 2002; Nordgren et al. 2016; Poole et al. 2008). Particulates become airborne from the movement of workers and animals to, from, and within animal housing facilities (Basinas et al. 2013; Gustafsson 1999), contributing to human exposures to bacteria, endotoxin, fungi (Douglas et al. 2018) viruses, dander, gases, and feed constituents (Duchaine et al. 2000). These airborne contaminants lead to a number of negative respiratory health outcomes including lung inflammation, airway hyper-responsiveness, and irritation of mucosal membranes (Charavaryamath et al. 2005; Cole et al. 2000; Senthilselvan et al. 2007). Even though researchers have been cataloging these harmful exposures for decades (*e.g*., Brouwer et al. 1986; Donham et al. 1977), they have not fully identified the riskiest contemporary work activities and the effectiveness of personal protective equipment (PPE) in this industry.

These knowledge gaps seriously limit the ability to propose suitable guidance for health protections among the estimated 31,000 IHO workers in the U.S. (BLS 2020). For example, in 2010, an expert panel concluded that researchers do not entirely understand which IHO workers could benefit most from the use of PPE (Von Essen et al. 2010). While we know that respirators can reduce the health effects from IHO contaminants (Dosman et al. 2000; Sundblad et al. 2006; Zejda et al. 1993), the panel was unable to determine whether PPE use for specific tasks is sufficient or whether PPE should be donned as soon as a worker enters an IHO; a potentially onerous and expensive recommendation that may not be feasible given harsh IHO work conditions.

A majority of data regarding health risks to U.S. agricultural workers comes from the Agricultural Health Study, a prospective cohort of over 50,000 North Carolinian and Iowan licensed pesticide applicators. While well-designed to study incident cancer outcomes related to pesticide exposures (Alavanja et al. 1996), and a follow-up interview was conducted to capture respiratory symptoms (Rinsky et al. 2019), it was not designed to focus specifically on IHO workers and therefore does not capture all farming activities. Further, this cohort consists largely of male managers (Alavanja et al. 1996), rather than those who work day-to-day inside concentrated animal feeding operation (CAFO) barns. Inclusion of both male and female workers in these studies is essential as there are known sex differences in respiratory outcomes among operation workers (Dosman et al. 2009, Senthilselvan et al. 2007).

One factor which restricts data collection on operations and limits to our understanding of IHO work conditions, exposures, and worker health outcomes is the blocking of access for researchers to CAFOs due to owner’s legal concerns. In addition, workers, who are often from marginalized communities including minorities, those with low-incomes, and lacking health insurance, may face job termination for participating in research efforts. This makes on-site air monitoring and the collection of health data exceedingly rare. Further, operations are often heterogenous in their age, design, animal density, animal life stages, and waste management systems. Therefore, comparing workers to one another from different operations without air sampling presents statistical problems and may lead to residual confounding due to differences in exposures (Radon et al. 2000), feed type (KimbellLDunn et al. 2001), barn construction (Kim et al. 2007), and activities between operations.

Fixed-effects regression analyses, which compare workers to themselves over time, can be used to examine exposure-outcome relationships and mitigate some of the threats to inference from heterogeneity in IHO sites. This technique has been successfully employed by Schinasi *et al*. who found strong associations between increasing CAFO odors and decrements in community health (Schinasi et al. 2011). To the best of our knowledge, no prior U.S. study has related self-reported work activities and PPE use to self-reported health outcomes among IHO workers who perform the day-to-day operations on industrial facilities. Further, no study of IHO worker activities and health outcomes has been analyzed using fixed-effects regression techniques. The purpose of this investigation was to identify factors that are associated with the respiratory and mucous membrane (eye, nose, and throat) health of the IHO workers we surveyed and to provide insight into factors for future research and interventions using an underemployed biostatistical method.

## METHODS

### Study population

In total, 103 IHO workers in North Carolina were recruited. Detailed methods on enrollment have been previously described (Nadimpalli et al. 2016). In brief, participants were recruited on a rolling basis from October 2013 to February 2014, with the last surveys completed in June 2014. Participants were eligible for inclusion in the sample population if they were: (1) current IHO workers (full- or part-time) and (2) agreed to participate in the study. Eligibility for inclusion in the baseline analysis population required that they provide survey data for the baseline enrollment visit. IHO workers were eligible for inclusion in the longitudinal analysis population if they were: (1) enrolled in the study and (2) completed at least one follow-up visit. Signed informed consent was obtained from each participant prior to participation. The study protocol was approved by the Johns Hopkins Bloomberg School of Public Health Institutional Review Board.

### Study location

North Carolina contains 10% of all pigs and hog operations in the U.S. and roughly 3,300 North Carolinian workers are employed in pig farming (NAICS code 1122) (BLS 2019). Located in southeast N.C., Duplin County is the second-greatest pork producing county in the U.S. (Food and Water Watch 2015). It is also home to the Rural Empowerment Association for Community Help (REACH; https://www.ncruralempowerment.org), who performed the recruitment, enrollment, and much of the data collection for this analysis.

### Questionnaires

#### Baseline

At enrollment, a baseline questionnaire was employed. It was designed to capture established work routines and health symptoms. Participants responded to survey questions consisting of how health, job tasks, and their work environment were “typically” or “usually” at their current IHO of employment. For example, participants were asked, “As part of your work, do you ever give antibiotics to pigs?” and “Do you usually have a cough?” Information about, and proxies for, the frequency, magnitude, and duration of contact with livestock, livestock manure, PPE use at work, animal species and life stage, typical job activities, and barn conditions were collected (see Supplemental Material, Questionnaires).

#### Follow-up

The follow-up questionnaire was adapted from the Agricultural Health Study (https://www.aghealth.nih.gov/collaboration/questionnaires.html), the American Thoracic Society (ATS-DLD-78-A), and Kimbell-Dunn *et al*. (KimbellLDunn et al. 1999). It was employed at two-week intervals for up to eight visits. Differing from the baseline questionnaire, it was designed to capture transient exposures and symptoms (see Supplemental Material, Questionnaires). Information about the frequency, magnitude, and duration of contact with pigs, job activities, personal behaviors (*e.g*., cigarette use), and PPE use at work was collected. Each question asked participants about the week prior to the study visit. For example, “In the past week have you…” In both questionnaires, with an attempt to capture a dose-response relationship, some questions asked participants to rate exposures using a Likert-like scale, while others asked for binary (ever/never or yes/no) responses.

### Statistical analyses

#### Baseline

At baseline, generalized logistic models clustered for household were used to assess the relationship between cross-sectional self-reported exposures and outcomes. Persons reporting at least one eye, nose, or throat symptom were grouped as mucous membrane cases. Due to collinearity, those who reported ever giving pigs shots and/or antibiotics were likewise grouped together. Biologically implausible values were dropped. Due to the small number of reported outcomes in some categories, analyses on exposures and outcomes with fewer than 5% of respondents indicating case status were not run.

*A priori* covariates explored in baseline analyses included age at enrollment (a continuous variable in years), sex (binary male/female), race/ethnicity (binary non-black Hispanic/other), asthma medication use if person reported being an asthmatic (binary controlled/uncontrolled), current smoking status (binary smoker/non-smoker), and season (summer, fall, winter, spring) (O’Shaughnessy et al. 2009), as well as days since last work shift (continuous). Based on prior knowledge and model fit, age and sex were included as baseline confounders in sensitivity analyses.

#### Follow-up

Longitudinal data were also checked for accuracy and variability. Exposures or outcomes with limited variability (≤ 1% of respondents) were *a priori* dropped from analyses to reduce any bias associated with small numbers (**Tables S4** and **S5**).

Scores were created for exposure activities that were similar in nature and displayed multi-collinearity (assessed via χ^2^ tests, with an α cutoff of 0.05). For example, poor *environmental barn conditions* consisted of reports of the following: vent fans turned off or non-existent at the facility (binary yes/no), extreme malodor (3 or 4 on a 4-point Likert-like scale), extreme temperature (3 or 4 on a 4-point Likert-like scale), a new herd entering the barns (binary yes/no), or extreme dust (3 or 4 on a 4-point Likert-like scale) in the past week. In main binary analyses, persons experiencing poor *environmental barn conditions* during that specific week were coded as a 1, whereas persons who reported none of the aforementioned activities were coded as referent 0. For sensitivity dose-response analyses, binary forms of each of the five input variable were summed; 1 being reported and 0 not being reported for that week, with a score of up to 5 for *environmental barn conditions*.

Using the same methodology, a *cleaning activity* score was created, consisting of on-IHO use of chemicals (binary yes/no), pressure washing the inside of barns (binary yes/no), application of pesticides (binary yes/no), and using a torch to clean the barns (binary yes/no) in the past week. In binary analyses, a person could have conducted a *cleaning activity* (coded as 1), or not (coded as 0), and in sensitivity trend analyses they could have a score of up to 4 reported activities.

In both the main binary *environmental barn conditions* and main binary *cleaning activity* analyses, summations of scores ≥ 2 were aggregated due to small numbers (1 of 711 for barn conditions and 12 of 738 for cleaning activities).

Intense *pig contact* activities (giving pigs medicine or shots) were also grouped using the same process, with main binary scores of 0 or 1 for each activity and sensitivity trend scores of 0 to 2.

Individual unweighted exposures and activities were also summed (1 as have been reported, and 0 if not reported in the past week) to assess the health implication for persons performing none to all 10 of the assessed activities. Up to six activities were reported in a single week by an individual, so scores of ≥ 4 were aggregated due to small numbers.

The use of PPE, including facemasks, eyewear, and full body suit/coveralls, was also grouped due to multi-collinearity. Participants were coded as a 1 in each category if they reported use of the specific PPE at least 80% of the time while at work in the past week and 0 if they reported using it less often. These scores were then summed, giving possible values from 0 to 3. Mask, eye protection, and coveralls were chosen for this analysis because we believed them to: (1) be *a priori* related to the outcomes of interest and (2) have variability in their use at baseline and over time and thus would not be dropped from fixed-effects regression models. For example, 726 of 737 (98.5%) of reported wearing boots 100% of the time at work in the past week; boot use was therefore not examined in models.

Reports of the number of times a person washed his/her hands was assessed in tertiles due to non-linearity. A report of handwashing 100 times per shift was dropped.

Groupings were also created for adverse health outcomes *a priori* based on biological understanding and number of case reports. These included reports of at least one respiratory symptom (*i.e*., excessive coughing, runny nose, difficulty breathing, or sore throat), at least one symptom that interfered with sleep (*i.e*., any symptoms reported, waking from sleep due to coughing, waking from sleep due to wheezing, or waking from sleep due to phlegm), sneezing, headache, and any reported mucous membrane symptom (*i.e*., eye or nose irritation).

Fixed-effects logistic regression was used to assess the relationship between self-reported exposures and outcomes in the past week and to control for time-invariant confounding variables (Allison 2009). These differences may include physical production facilities (*i.e*., number and life stage of pigs or waste management systems) and the operational structures (*i.e*., how often pits under slatted floors are drained). Confounders of interest from the literature and relevant to the longitudinal fixed-effects analysis included only month of follow-up visit (Basinas et al. 2013).

All data were analyzed using Stata (StataCorp. 2017. *Stata* Statistical Software: Release 15. College Station, TX: *StataCorp* LP).

## RESULTS

In this study of IHO workers, elevated odds of self-reported symptoms by those who ever performed activities and those who performed increasingly hazardous transient activities on the IHO were found. Due to small case numbers, confidence intervals are often wide, but effect estimates are large and demonstrate a consistency in magnitude and direction across main and sensitivity analyses.

At baseline, 103 current workers entered the cohort through rolling admissions. As reported in previous studies, these workers (with 1-27 years of IHO experience) were primarily non-black Hispanic (88%), male (55%), and aged 16-62 years (Nadimpalli et al. 2016). Most did not live on the same property as an IHO (92%) (**Table 1**). An average of 8 years working on any IHO was reported, as was an average of 6.4 days worked per week and a majority of work time spent in direct contact with pigs (82%) (**Table 2**). The most prevalent work activities employees reported *ever* performing included handling dead pigs (79%), giving pigs shots or injections (69%), having direct contact with pig manure (67%), administering antibiotics (62%), applying pesticides in or around barns (49%), and drawing blood from pigs (9%). Participants were also asked to classify typical mask usage at work, and 38% responded that they always wore a mask (**Table 2**).

**Table 1.** Baseline demographic and household characteristics of the industrial hog operation worker cohort, North Carolina, 2013-2014.

| Characteristic | Reports |
| --- | --- |
| Workers in cohort, *n* | 103 |
| Age in years, *mean ± SD* | 38 ± 11 |
| Sex, *n (%)* |  |
| Male | 55 (54) |
| Female | 46 (46) |
| Race/ethnicity, *n (%)* |  |
| Hispanic, non-black | 88 (88) |
| Black | 12 (12) |
| Education status, *n (%)* |  |
| Less than high school education | 47 (47) |
| High school degree/GED or higher | 52 (53) |
| Body mass index (BMI), *n (%)* |  |
| <30.0 | 58 (56) |
| ≥30.0 | 38 (37) |
| Used a gym or workout facility in the last three months, *n (%)* |  |
| Yes | 9 (9) |
| No | 92 (91) |
| Current cigarette smoker, *n (%)* |  |
| Yes | 13 (17) |
| No | 65 (83) |
| Health insurance, *n (%)* |  |
| Yes | 48 (48) |
| No | 52 (52) |
| Place where IHO workers seek medical care,<sup>a</sup> *n (%)* |  |
| Private doctor | 49 (49) |
| Emergency department or urgent care center | 29 (28) |
| Hospital | 18 (17) |
| Free clinic | 16 (16) |
| Other | 3 (3) |
| Does not seek medical care under any circumstance | 4 (4) |
| Had a cat or dog, *n (%)* |  |
| Yes | 44 (43) |
| No | 50 (47) |
| Lived on same property as an IHO, *n (%)* |  |
| Yes | 8 (8) |
| No | 89 (92) |
| Month of baseline visit, *n (%)* |  |
| January | 1 (1) |
| February | 50 (49) |
| October | 30 (29) |
| November | 22 (21) |

Note. IHO = industrial hog operation.
a. Categories are not mutually exclusive.

**Table 2.** Baseline self-reported occupational exposure activities among industrial hog operation workers, North Carolina, 2013-2014.

| Characteristic | Reports |
| --- | --- |
| Years worked on any IHO, *mean ± SD* | 8 ± 6 |
| Days worked per week, *mean ± SD* | 6.4 ± 0.8 |
| Percent of time at work spent in direct contact with pigs, *mean ± SD* | 82 ± 27 |
| Ever handled dead pigs, *n (%)* | 77 (79) |
| Ever gave pigs shots or injections, *n (%)* | 68 (69) |
| Ever came into direct contact with or touched pig manure, *n (%)* | 61 (67) |
| Ever gave antibiotics to pigs, *n (%)* | 60 (62) |
| Ever drew blood or collect other fluids from pigs, *n (%)*, *n (%)* | 9 (9) |
| Only worked with |  |
| Sows, nursery, and/or weaning pigs | 46 (48) |
| Feeder and/or finisher pigs | 24 (25) |
| Ever applied pesticides inside or around the barns, *n (%)* | 48 (49) |
| Wore coveralls/full body suit, *n (%)* |  |
| Always | 68 (70) |
| Sometimes | 14 (14) |
| Never | 15 (16) |
| Wore a mask, *n (%)* |  |
| Always | 37 (38) |
| Sometimes | 43 (44) |
| Never | 18 (18) |
| Wore glasses/goggles, *n (%)* |  |
| Always | 22 (22) |
| Sometimes | 34 (35) |
| Never | 42 (43) |
| Work clothes ever washed with the laundry of household members, *n (%)* | 16 (17) |

Participants were asked whether they ever experienced a variety of symptoms, outside of having a cold or the flu. Respondents most frequently reported having eye irritation (18%), nose irritation (16%), throat irritation (15%), any allergies (13%), and doctor-diagnosed asthma (9%; **Table 3**).

**Table 3.** Baseline self-reported health conditions among industrial hog operation (IHO) workers, 679 North Carolina, 2013-2014.

| Characteristic<sup>a</sup> | Prevalence, *n* (%) |
| --- | --- |
| Ever had eye irritation | 19 (19) |
| Within the last month | 12 (63) |
| Ever had nose irritation | 16 (16) |
| Within the last month | 10 (67) |
| Ever had throat irritation | 15 (15) |
| Within the last month | 13 (87) |
| Any allergies | 13 (13) |
| Doctor-diagnosed asthma | 9 (9) |

Note. *a.* Participants were asked to not report health outcomes that they attributed to having a cold.

Those who reported *ever* drawing pig blood had an increased likelihood of reporting eye, nose, and throat symptoms (PR: 3.7; 95% confidence interval (CI): 1.9, 7.0) and any allergies (PR: 4.4; 95% CI: 1.7, 12) (**Table 4**). Increased prevalence of eye, nose, or throat symptoms were also reported in those who ever applied pesticides in or around pig barns (PR: 2.2; 95% CI: 1.0, 4.8) and those who washed work clothes with household laundry (PR: 2.3; 95% CI: 1.0, 5.3). Across tertiles of years worked on any IHO, increasing eye, nose, and throat symptoms were reported (*p*-for trend: 0.01), and in the uppermost tertile of exposure (PR: 4.3; 95% CI: 1.5, 12). This trend was also seen in the association between eye, nose, and throat symptoms and tertiles of percent of life worked on any IHO (*p*-for trend: 0.04; highest tertile PR: 3.4; 95% CI: 0.99, 11) (**Table 4**).

**Table 4.** Crude baseline associations between binary (unless noted as tertiles) self-reported industrial hog operation (IHO) work activities and binary self-reported symptoms among IHO workers, North Carolina, 2013-2014, clustered at the household level.

| Reported characteristic | Eye, nose, or throat symptoms |  |  | Any allergies |  |  | Doctor-diagnosed asthma |  |  |
| --- | --- | --- | --- | --- | --- | --- | --- | --- | --- |
|  | *n* | PR (95% CI) | *p*-value | *n* | PR (95% CI) | *p*-value | *n* | PR (95% CI) | *p*-value |
| Have you ever |  |  |  |  |  |  |  |  |  |
| Given pigs shots and/or antibiotics | 96 | 2.2 (0.8, 6.6) |  | 97 | 0.6 (0.2, 1.5) |  | 97 | 1.0 (0.2, 5.2) |  |
| Drawn pig's blood | 97 | 3.7 (1.9, 7.0) |  | 98 | 4.4 (1.7, 12) |  | 98 | 3.3 (0.9, 12) |  |
| Handled pig manure | 90 | 1.0 (0.4, 2.4) |  | 91 | 1.3 (0.4, 4.6) |  | 91 | 1.5 (0.3, 7.0) |  |
| Applied pesticides in or around the barns | 97 | 2.2 (1.0, 4.8) |  | 98 | 2.3 (0.8, 6.9) |  | 98 | 1.0 (0.2, 4.7) |  |
| Washed work clothes with household laundry | 95 | 2.3 (1.0, 5.3) |  | 96 | 0.5 (0.1, 3.3) |  | 96 | 0.7 (0.1, 5.7) |  |
| Do you typically |  |  |  |  |  |  |  |  |  |
| Work exclusively in sow, nursery, and/or farrow barns | 93 | 0.8 (0.3, 1.7) |  | 94 | 0.5 (0.2, 1.6) |  | 94 | 2.1 (0.4, 11) |  |
| Work exclusively in feeder and/or finisher barns | 93 | 1.3 (0.5, 3.1) |  | 94 | 1.0 (0.3, 3.4) |  | - | - |  |
| Always wear coveralls/full body suit, mask, and eye protection | 96 | 1.3 (0.5, 3.4) |  | 97 | 3.8 (1.4, 9.9) |  | 97 | 2.6 (0.6, 12) |  |
| Work 7 days per week | 96 | 0.4 (0.1, 1.1) |  | 97 | 0.1 (0.01, 0.6) |  | 97 | 0.3 (0.1, 1.7) |  |
| Years worked on any IHO | 85 |  |  | 85 |  |  | 87 |  |  |
| Tertile 1 (1-5 years) |  | Ref (1.0) |  |  | Ref (1.0) |  |  | Ref (1.0) |  |
| Tertile 2 (6-10 years) |  | 1.9 (0.5, 7.6) |  |  | 0.6 (0.1, 2.6) |  |  | 3.6 (0.4, 35) |  |
| Tertile 3 (11-27 years) |  | 4.3 (1.5, 12) |  |  | 0.8 (0.2, 2.7) |  |  | 3.0 (0.3, 30) |  |
| Trend |  |  | 0.01 |  |  | 0.6 |  |  | 0.3 |
| Percent of life working on any IHO | 82 |  |  | 83 |  |  | 55 |  |  |
| Tertile 1 (2.4-11.6%) |  | Ref (1.0) |  |  | Ref (1.0) |  |  | Ref (1.0) |  |
| Tertile 2 (11.7-26.3%) |  | 1.8 (0.4, 7.2) |  |  | 1.04 (0.3, 3.8) |  |  | 1.5 (0.3, 8.4) |  |
| Tertile 3 (26.4-51.9%) |  | 3.4 (0.99, 11) |  |  | 0.84 (0.2, 3.5) |  |  | - |  |
| Trend |  |  | 0.04 |  |  | 0.8 |  |  | 0.1 |

Note. PR = prevalence ratio. CI = confidence interval. - = model did not converge. IHO = Industrial hog operation. Crude models are presented as main tables in this analysis due to convergence issues in adjusted models and the consistency between point estimates in crude and adjusted regressions. In alternative sensitivity analyses (not shown) prevalence odds ratios (PORs) were calculated and showed to overestimate the exposure-response association. Due to this, prevalence ratios (PRs) were presented for baseline analyses.

In the unhypothesized direction, reports of always wearing all three PPE (full body suit/coveralls, mask, and eye protection) on the job were associated with higher prevalence of reports of any allergies (PR: 3.8; 95% CI: 1.4, 9.9), while working all seven days per week compared to those working less often was associated with lower prevalence of allergies (PR: 0.1; 95% CI: 0.01, 0.6) (**Table 4**). In models that converged, these associations were consistent in direction and magnitude after adjustment for age and sex (**Table S1**).

Of the 101 persons eligible for longitudinal data analyses, 95% of their study visits were completed and 90 of the 101 participants completed all eight follow-up visits (**Figure 1**). Multiple imputation was not conducted as, overall, very few data points were missing (∼5-10% per analysis) and the missingness was determined to not be at random.

**Figure 1.**
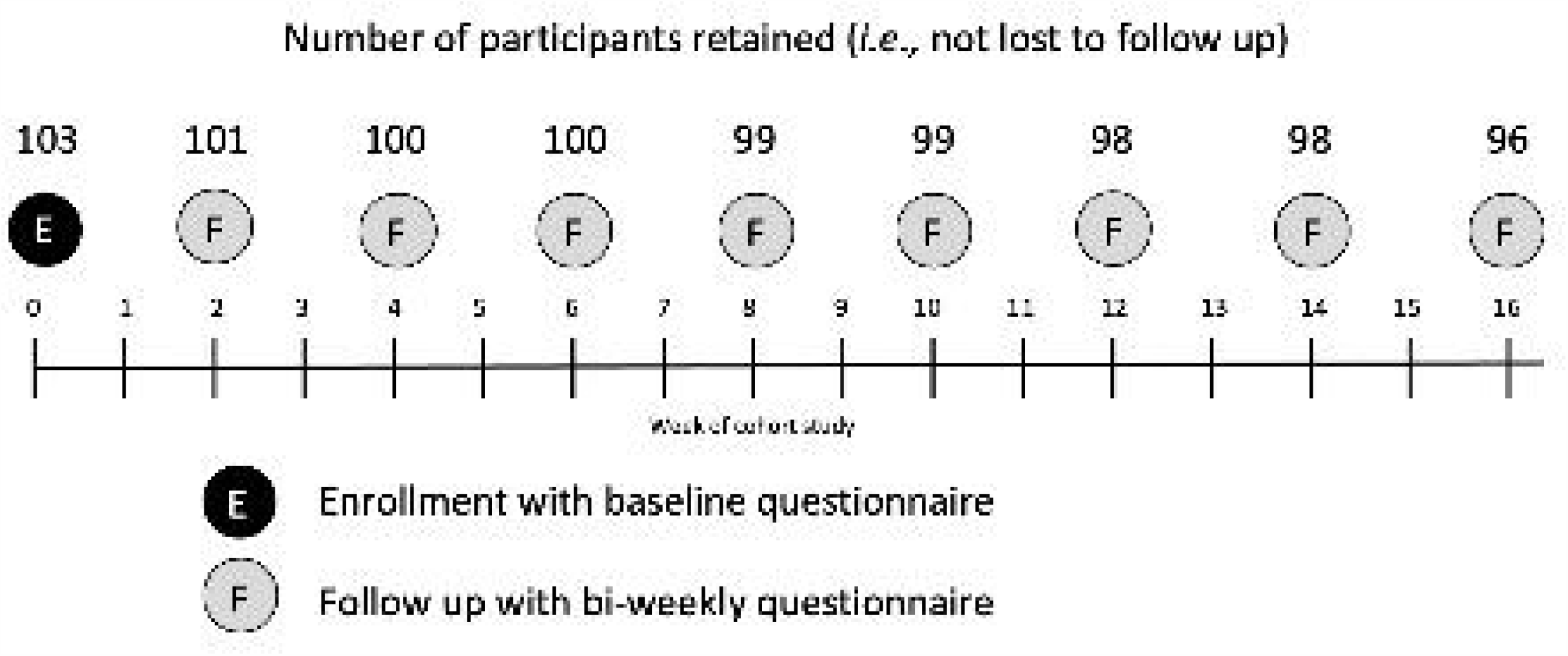
Sampling scheme and loss-to-follow-up between the baseline and bi-weekly study visits within a cohort of industrial hog operation (IHO) workers, North Carolina, 2013-2014.

During the previous week, persons reported working an average of 6 ± 1 days per week, 42 ± 12 hours per week, with 38 ± 14 hours in direct contact with pigs. High-frequency work activities included administering shots (49%) and using cleaning chemical(s) (56%) (**Table 5**).

**Table 5.** Time-varying occupational exposure activities occurring during the week immediately preceding the biweekly study visit among industrial hog operation workers, North Carolina, 2013-2014.

| Activities in the past week | Affirmative responses |
| --- | --- |
| Number of days worked, *mean ± SD* | 6 ± 1 |
| Number of hours worked, *mean ± SD* | 42 ± 12 |
| Number of hours in direct contact, *mean ± SD* | 38 ± 14 |
| Number of sick pigs, *mean ± SD* | 61 ± 166 |
| Number of dead pigs, *mean ± SD* | 42 ± 120 |
| % of time coveralls/full body suit were worn, *mean ± SD* | 81 ± 38 |
| % of time a mask was used, *mean ± SD* | 54 ± 46 |
| % of time eye protection used, *mean ± SD* | 28 ± 42 |
| Number of times washed hands at the IHO, *mean ± SD* | 8 ± 6 |
| Barn condition score factors, *n (%)* |  |
| Vent fans were off | 178 (34) |
| Malodor |  |
| None, moderate | 564 (76) |
| Extreme | 175 (24) |
| Temperature in the barns, *n (%)* |  |
| Cold, comfortable | 614 (85) |
| Hot | 111 (15) |
| A new herd entered the barn(s), *n (%)* | 47 (6) |
| Dustiness in barns, *n (%)* |  |
| None, moderate | 705 (96) |
| Extreme | 32 (4) |
| Cleaning and pesticide score factors, *n (%)* |  |
| Used cleaning chemical(s) at the IHO | 414 (56) |
| Pressure washed | 290 (39) |
| Applied pesticides | 224 (30) |
| Used a torch | 20 (3) |
| Pig contact score factors, *n (%)* |  |
| Gave pigs medicine | 241 (68) |
| Gave pigs shots | 363 (49) |
| Received an influenza vaccine since the last study visit, *n (%)* | 21 (3) |

Note. SD = standard deviation. IHO = industrial hog operation.

Outcomes of high prevalence and variability reported in the same bi-weekly surveys included any respiratory health symptom (6%), consisting of excessive coughing (3%), runny nose (3%), difficulty breathing (2%), sore throat (1%), shortness of breath (1%), wheezing or whistling in chest (1%), and/or chest tightness (0%, 2 cases). Symptoms interfering with sleep were reported in 3% of surveys and sneezing and headache in 2% of surveys. Mucous membrane symptoms, consisting of burning, tearing, or irritated eyes (1%) and/or burning or irritated nose (1%), were also reported in 2% of bi-weekly surveys (**Table 6**).

**Table 6.**
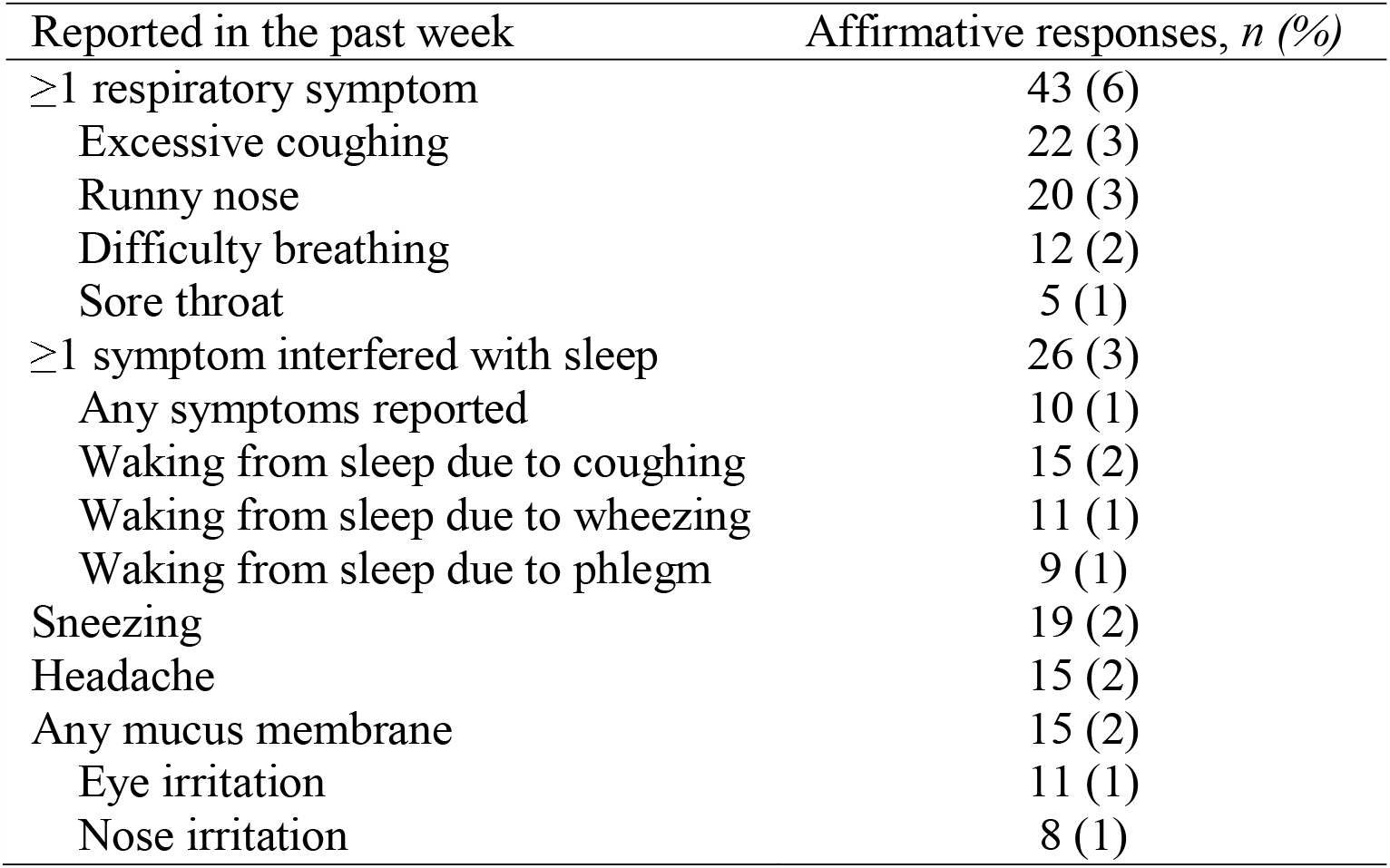
Time-varying acute health outcomes occurring during the week immediately preceding the biweekly study visit among industrial hog operation workers, North Carolina, 2013-2014.

Due to concerns about false reporting of symptoms or work conditions, researchers included ten “dummy” symptoms (*i.e*., without any known prior association with exposure to hog production facilities, such as difficulty urinating) on the questionnaires. None of the dummy symptoms were reported more than six times within the 752 person-records.

Time-varying acute health outcomes and work activities were assessed using fixed-effects regression to control for time-invariant confounders. Consistency was seen between an increased risk of reported symptoms during weeks when workers engaged in activities that produced or retained dust within barns, conducted cleaning activities, and engaged in activities that involved close contact with pigs (13 of 14). Administering pigs medicine or shots was the riskiest activity category for all symptoms examined. Higher odds of headache, respiratory symptoms, and symptoms interfering with sleep were observed during weeks when workers conducted two or more categories of activities compared to zero or one. Also, as hypothesized, a protective effect was estimated for any PPE use (compared to when none was used) in all five outcomes. Handwashing was also protective in 4 of 5 outcomes during weeks when done at least eight times per shift (the median) versus fewer times (**Tables 7** and **8**).

**Table 7.** Crude time-varying acute health outcomes and binary work activities the week immediately preceding the biweekly study visit among industrial hog operation workers, North Carolina, 2013-2014.

| Reported in the past week | At least one respiratory symptom<sup>a</sup> |  | At least one symptom interfered with sleep<sup>b</sup> |  |
| --- | --- | --- | --- | --- |
|  | obs. (groups) | OR (95% CI) | obs. (groups) | OR (95% CI) |
| Any hot or dusty barn conditions<sup>c</sup> | 225 (31) | 4.0 (1.4, 12) | 152 (21) | 1.7 (0.5, 5.3) |
| Conducted any pesticide application or cleaning activity<sup>d</sup> | 229 (31) | 3.0 (1.2, 7.5) | 147 (20) | 4.5 (1.1, 18) |
| Administered pigs medicine or shots<sup>e</sup> | 223 (30) | 6.8 (1.8, 25) | - | - |
| Two or three of the above categories<sup>f</sup> | 215 (30) | 10 (2.2, 46) | 144 (20) | 19 (2.1, 171) |
| Used any PPE<sup>g</sup> | 226 (31) | 0.3 (0.1, 1.5) | 154 (21) | 0.1 (0.01, 0.8) |
| Washed hands at least 8 times per shift<sup>h</sup> | 228 (31) | 0.3 (0.1, 0.8) | 147 (20) | 0.6 (0.2, 1.8) |

Note. OR = odds ratio. CI = confidence interval. - = model did not converge. PPE = personal protective equipment. OR (95% CI) estimates are derived from conditional logistic fixed-effects regression models, which estimate the average of all within-person differences between time-varying exposure and outcome.
a. Excessive coughing, runny nose, difficulty breathing, or sore throat.
b. Any sleep symptoms reported, waking from sleep due to coughing, waking from sleep due to wheezing, or waking from sleep due to phlegm.
c. Sum of vents off (yes=1, no=0), extreme malodor (yes=1, no=0), hot temperature (yes=1, no=0), a new herd entering the barn(s) (yes=1, no=0), and extreme dust (yes=1, no=0)
d. Sum of used cleaning chemicals (yes=1, no=0), pressure washed (yes=1, no=0), used pesticides (yes=1, no=0), and used a torch (yes=1, no=0)
e. Sum of gave pigs medicine (yes=1, no=0) and gave pigs shots (yes=1, no=0)
f. Number of individual activities/conditions (maximum 10)
g. Sum of consistently (≥80% of the time at work) wore the following: coveralls/full body suit (yes=1, no=0), mask (yes=1, no=0), and glasses (yes=1, no=0).
h. 8 is the median number of times workers reported washing their hands per IHO work shift.

**Table 8.** Crude time-varying acute health outcomes and binary work activities the week immediately preceding the biweekly study visit among industrial hog operation workers, North Carolina, 2013-2014.

| Reported in the past week | Sneezing |  | Headache |  | Eye or nose irritation |  |
| --- | --- | --- | --- | --- | --- | --- |
|  | obs.<br>(groups) | OR (95% CI) | obs.<br>(groups) | OR (95% CI) | obs.<br>(groups) | OR (95% CI) |
| Any hot or dusty barn conditions<sup>a</sup> | 85 (12) | 0.8 (0.2, 3.3) | 92 (12) | 2.6 (0.8, 9.0) | 84 (11) | 8.4 (1.0, 71) |
| Conducted any pesticide application or cleaning activity<sup>b</sup> | 86 (12) | 3.5 (0.97, 12) | 93 (12) | 1.9 (0.5, 7.1) | 85 (11) | 2.6 (0.6, 11) |
| Administered pigs medicine or shots<sup>c</sup> | 87 (12) | 12 (1.3, 105) | 93 (12) | 4.6 (0.8, 25) | 70 (9) | 15 (1.7, 133) |
| Two or three of the above categories<sup>d</sup> | 85 (12) | 4.1 (1.1, 16) | 92 (12) | 7.5 (1.4, 40) | 67 (9) | 6.1 (1.3, 28) |
| Used any PPE<sup>e</sup> | 85 (12) | 0.1 (0.01, 1.0) | 90 (12) | 0.9 (0.1, 6.2) | 82 (11) | 0.1 (0.02, 0.9) |
| Washed hands at least 8 times per shift<sup>f</sup> | 87 (12) | 0.9 (0.24, 3.2) | 93 (12) | 1.3 (0.3, 5.4) | 85 (11) | 0.6 (0.2, 2.2) |

Note. OR = odds ratio. CI = confidence interval. PE = personal protective equipment. OR (95% CI) estimates are derived from conditional logistic fixed-effects regression models, which estimate the average of all within-person differences between time-varying exposure and outcome.
*a.* Sum of vents off (yes=1, no=0), extreme malodor (yes=1, no=0), hot temperature (yes=1, no=0), a new herd entering the barn(s) (yes=1, no=0), and extreme dust (yes=1, no=0)
*b.* Sum of used cleaning chemicals (yes=1, no=0), pressure washed (yes=1, no=0), used pesticides (yes=1, no=0), and used a torch (yes=1, no=0)
*c.* Sum of gave pigs medicine (yes=1, no=0) and gave pigs shots (yes=1, no=0)
*d.* Number of individual activities/conditions (maximum 10)
*e.* Sum of consistently (≥80% of the time at work) wore the following: coveralls/full body suit (yes=1, no=0), mask (yes=1, no=0), and glasses (yes=1, no=0)
*f.* 8 is the median number of times workers reported washing their hands per IHO work shift

In a sensitivity analysis, exposures were modeled via scores in an effort to assess a dose-response (**Tables S2** and **S3**). Higher odds of reporting health impacts were observed with increasing exposures and worsening work conditions. The most profound effects were seen in the upper ends of exposure categories, with a *p*-for-trend <0.05 for 11 of 25 associations. The use of PPE showed a protective effect in 14 of 15 outcome groups with a no-threshold effect (**Table S2**). Sensitivity analyses that included the addition of a calendar month dummy variable were also performed. In those models which converged, estimates of similar magnitude and direction were observed (data not shown).

## DISCUSSION

In this study of self-reported IHO work activities and health outcomes, we found that during the weeks when workers performed the most unfavorable job tasks they also experienced increased odds of respiratory and mucous membrane irritation symptoms. In addition, they reported reduced odds of symptoms when they wore PPE or washed their hands more frequently. To the best of our knowledge, our study is the first to apply fixed-effects regression as a tool to measure associations between self-reported workplace exposures and health outcomes in an IHO worker cohort. In addition, our study is more generalizable to day-to-day workers than prior work, where most cohort participants have been white male farm supervisors not in direct contact with swine (Alavanja et al. 1996). For example, in Iowa, the nation’s largest producer of swine, Donham *et al*. enrolled 2,059 hog operation owners with five years of follow-up and three annual visits (Donham et al. 1990). From this cohort, 40 operations were determined to be CAFOs with 207 IHO workers enrolled. Of the 207 IHO workers, 100% were white, 88% were male, and 20% were smokers, limiting generalizability to a workforce who, in practice, is often made of non-white immigrants and, in our study, 46% female.

As in our study, Donham *et al*. did find that participants who worked in IHOs (compared to those working at non-confinement operations) had more chronic and acute respiratory health symptoms (Donham et al. 1990). Unlike our study, in which we did not attempt to gain access on-IHO, Donham *et al*. benefitted from on-operation collection of ambient air samples while workers worked and samples from inside their masks; however, they failed to collect data regarding specific work activities.

In congruence with the Agricultural Health Study (Alavanja et al. 1996), our cohort consisted primarily of non-smokers; however, our participants had a higher prevalence of self-reported asthma (8.7%) than those in the Agricultural Health Study (5.1%) (Henneberger et al. 2014). The prevalence of asthma in our predominantly non-black Hispanic adult population was also higher than national average for non-black Hispanic adults (6.4%) (Moorman et al. 2011). While capturing the incident development of asthma in our study was not possible, we did observe an association in increased reported cases of doctor-diagnosed asthma with increasing tertiles of years worked on any IHO (PR: 2.6; 95% CI: 1.03, 6.3), which is consistent with prior reports of increasing trends in development of asthma among farmers (Holness et al. 1995).

As hypothesized, reports of ever drawing pig blood, applying pesticides, and increasing years worked at any IHO were consistently associated with increased reports of eye, nose, or throat symptoms. Acute exposures, including those associated with dustiness and cleaning of barns and close contact with pigs were associated with increased odds of a variety of symptoms, particularly in the highest categories of exposure. Completing more of these tasks showed an increase in symptom reporting as well, evidence for a need to rotate job tasks among employees and to create work environments that are inherently less dusty.

Unexpectedly, working an average of seven days per week was associated with decreased reports of symptoms. This association is a potential indication of healthy worker effect bias where only the healthiest and most tolerant of symptoms report for work every day. It may also be due to the fact that when workers are away from work exposures their respiratory system rebounds, leading to inflammatory reactions (Bønløkke et al. 2012). Another unexpected finding was that at baseline, workers who reported “always” wearing all three types of PPE at work reported increased odds of allergies. This is potentially a case of reverse causation or reporting errors, as this finding was not corroborated in our longitudinal analyses.

In weeks when wearing PPE and washing hands at or above the median frequency, workers had decreased odds of symptoms compared to weeks when they did not wear PPE or when they washed their hands less frequently. It has been shown that exposure to pesticides and other respiratory irritants can be modified by the use of PPE (Salvatore et al. 2008). In particular, N95 masks have been shown to block harmful pathogens found on IHOs (Ferguson et al. 2014).

While IHO workers in our study were not queried directly on which masks were used, subsequent assessment of a small number of different workers in a follow-up study (*n*=18) suggested that most adult IHO workers wear an employer-provided N95 respirator (15/17) and, less commonly, a surgical mask (2/17) (Coffman 2018). Of the 17 whose employer provided them a mask, 16 did so in the past two weeks, and all 17 reported using the mask provided (Coffman 2018). Employer training in face mask usage was also high (15/19) (Coffman 2018), which may partly explain the rate of use in this pilot study. Ferguson *et al*. have also documented that IHO workers are willing to become educated in personal protection and documented that they found value in learning about methods to protect themselves from exposures (Ferguson et al. 1989). Intervention studies that examine employer-provided handwashing stations and increased access to PPE are areas of future interest.

### Limitations

Several limitations potentially temper the current analysis. Because recruitment was non-random, participants represent a self-selected group, which may lead to potential selection bias; including those who may be more concerned about work practices, are sicker than those who did not participate, and those who are willing to be involved in a study (which may have resulted in termination from their job had their employer discovered their participation). That said, the recruited population reflects the occupational demographics of the area of southeastern North Carolina. Second, in order to maintain worker confidentiality, operations could not be accessed directly, and therefore air sampling and personal monitoring could not be conducted on-site to corroborate survey responses. Third, baseline analyses lack needed temporality to make conclusions regarding causality and the small sample size (*n*=103) of our baseline analyses make the results highly sensitive to outliers. Also, because of small numbers of varying reports of exposures or outcomes between weeks, the confidence interval for main and trend analyses are wide. However, the number, magnitude, and direction of estimates of association demonstrate strong consistency, which indicates that work on IHOs is detrimental to physical health. Finally, these data are from IHO workers, who may represent a healthy-worker population, but this bias would most likely drive associations toward to the null.

### Strengths

Fixed-effects regression was used to eliminate data collection on the vast array of unmeasurable confounders that arise from a lack of access to IHOs and from differences due to between-person perception of magnitude of symptoms or work conditions. The use of fixed effects modeling also controls for characteristics on IHOs that do not change temporally during our study, such as feed type or barn construction (*e.g*., floor slatting). Additionally, this modeling technique partially ameliorated challenges with model convergence, given a number of low-prevalence confounders. Another strength of our study was the ability to recruit day-to-day IHO workers who are typically nervous about participating in research due to potential employer backlash. By working in tandem with a local organization who has strong ties to the community we were able to establish trust in our data security and provide laborers with information which may help them protect their health in the future.

## Conclusions

In this analysis of self-reported work activities and health among 103 IHO employees, we observed positive associations between exposures and proxies for exposures to possible respiratory irritants and reported respiratory health and mucous membrane outcomes, using each participant as their own control. Fixed-effect regression analysis of repeated measures differs from prior research using traditional between-person estimates. Our study also differed as we recruited of day-to-day workers and women leading to a more representative sample of the in-barn workforce. Our data show that when exposed to unfavorable tasks the risk of mucous membrane and respiratory symptoms increased, while when handwashing increased or PPE was employed, the risk of reporting negative health outcomes was reduced. Further research should focus on what types of PPE are most appropriate and functional in this workplace environment and employers may wish to focus on activities that increase job rotation, decrease dust exposure, and provide adequate access to handwashing stations and PPE.

## Data Availability

Due to the exceedingly sensitive nature of the data collected, it will not be made publicly available.

## ACKNOWLEDGEMENTS

This study would not have been possible without a strong partnership between researchers and North Carolinian community-based organization members who have fostered the trust of too often marginalized and at-risk community members. The authors deeply thank the workers who participated in this study.

This manuscript is dedicated to the memory of Dr. Steve Wing, who helped conceive the design and analytical framework for this cohort study.

## Notes

### Competing Interest Statement

The authors have declared no competing interest.

### Funding Statement

Funding for this study was provided by National Institute for Occupational Safety and Health (NIOSH) grant K01OH010193; Johns Hopkins NIOSH Education and Research Center grant T42OH008428; a directed research award from the Johns Hopkins Center for a Livable Future; the Johns Hopkins NIOSH Education and Research Center Pilot Award; award 018HEA2013 from the Sherrilyn and Ken Fisher Center for Environmental Infectious Diseases Discovery Program at the Johns Hopkins University, School of Medicine, Department of Medicine, Division of Infectious Diseases; and National Science Foundation (NSF) grant 1316318 as part of the joint NSF National Institutes of Health (NIH) U.S. Department of Agriculture Ecology and Evolution of Infectious Diseases program. V.R.C was supported by the Johns Hopkins Center for a Livable Future-Lerner Fellowship. N.P. was supported by NIH/National Institute of Environmental Health Sciences (NIEHS) grant 5T32ES007141. D.L was supported by funds from the Johns Hopkins Center for a Livable Future. M.N. was supported by a Royster Society fellowship and a U.S. Environmental Protection Agency Science to Achieve Results fellowship. C.D.H. was supported by NIOSH grant K01OH010193, E.W. Al Thrasher Award 10287, NIEHS grant R01ES026973, and NSF grant 1316318. The funders had no role in study design, data collection and analysis, decision to publish, or preparation of the manuscript.

### Author Declarations

The study protocol was approved by the Johns Hopkins Bloomberg School of Public Health Institutional Review Board.

